# Direct Fast Scarlet and Congo Red Staining in Identification of Eosinophils and their Cell-free Granules in Cardiac Tissue

**DOI:** 10.1101/2024.12.25.24319622

**Authors:** Takayuki Kumaki, Tomoe Abe, Takeshi Kashimura, Shigeharu Ueki, Hajime Umezu, Souma Sato, Shou Hirayama, Hayao Ikesugi, Kazuyo Tanaka, Yuka Sekiya, Hiroki Tsuchiya, Ryohei Sakai, Hiromi Kayamori, Shinya Fujiki, Tsugumi Takayama, Takayuki Inomata

**Author notes:** Correspondence to: Takeshi Kashimura, MD, Department of Cardiovascular Medicine, Niigata University Graduate School of Medical and Dental Sciences, 1-757 Asahimachidori, Chuo-ku, Niigata, 951-8510, Japan.

## Abstract

**Background:** Cell-free eosinophil granules, which are considered harmful to the heart, are stained red by haematoxylin-eosin (HE); however, they can be overlooked in cardiac tissue due to the reddish staining of cardiomyocytes and fibres. Direct fast scarlet (DFS) and Congo red (CR), known for staining amyloid deposits, stain eosinophil granules; however, no firm evidence currently exists. This study aimed to confirm that DFS and CR stain eosinophil granules red and evaluate their advantages over HE.

**Methods:** Paraffin-embedded endomyocardial biopsy samples from six patients, each with eosinophil-infiltrating cardiac disorders, and six patients with lymphocytic myocarditis were stained.

**Results:** The distributions of red granules stained with DFS and CR were similar to those of red granules stained with HE in serial sections. Major basic protein (MBP), a marker of eosinophil granules, was detected within cells positive for galectin-10, a marker of eosinophil cytoplasm. These MBP-positive granules, pre-scanned using immunofluorescence, were stained with a reddish hue by HE, DFS, and CR. MBP-positive granules surrounding galectin-10-negative cells with a degenerated nucleus, characteristic of cytolytic eosinophil degranulation (ETosis), were identified by HE, DFS, and CR staining. Non-granular MBP-positive interstitial areas were not identified by HE, DFS, or CR staining, suggesting that these staining methods did not detect deposited granule proteins released from disrupted eosinophil granules. Eosinophil granules were identified by extracting the red colour using Image-J software in DFS-stained images, more specifically than in CR-stained images and not in HE-stained images. Cardiologists counted more eosinophils in DFS-stained sections than in HE-stained serial sections within a certain time without miscounting.

**Conclusion:** Our results demonstrated the potential of DFS as a superior method for identifying eosinophils and their cell-free granules in cardiac tissues. DFS may enhance the detection of eosinophils and improve the treatment of eosinophil-related heart diseases.

**KEY MESSAGES:**

- **What is already known on this topic** – Several case reports have demonstrated that red-stained granules observed in direct fast scarlet (DFS)- or Congo red (CR)-stained sections exhibit distribution patterns similar to those of eosinophil granules in haematoxylin-eosin (HE)-stained serial sections.
- **What this study adds** – This study provides the first direct evidence that red-stained granules observed with DFS and CR staining are eosinophil granules, as shown by the presence of major basic protein (MBP). Further, this study demonstrated that identifying eosinophil granules is most effective with DFS staining, which, compared to HE staining, allowed a more rapid counting of eosinophils by non-pathologist cardiologists without miscounts.
- **How this study might affect research, practice or policy** – This study may enhance the accuracy of eosinophil count and cytolytic degranulation detection, potentially via machine counting, and improve quantitative and qualitative definitions of eosinophil-infiltrating heart diseases.

## INTRODUCTION

Eosinophil infiltration and the release of specific granule proteins have long been associated with cardiac tissue injury and impaired cardiac function in heart disease,^1–3^ and histological diagnosis is essential.^4,5^ Eosinophils are identified in cardiac tissue by haematoxylin-eosin (HE) staining, typically as small spherical cells with a bi-lobed nucleus and multiple bright red cytosolic granules.^6^ However, because eosin also stains cardiomyocytes and other fibres pink, distinguishing eosinophils can be challenging. Immunohistochemistry for eosinophil-specific molecules is reliable;^6^ however, it can be time-consuming and costly. Therefore, a simple, inexpensive, and reliable method for eosinophil detection in cardiac tissue is necessary to avoid their misidentification.

Eosinophils have approximately 200 eosinophilic granules within the cell, which contain highly cytotoxic cationic proteins, such as major basic protein (MBP).^6^ The release of granule proteins (degranulation) from vital eosinophils takes the form of piecemeal degranulation or exocytosis.^7^ Additionally, a cell death-mediated degranulation mechanism, called cytolysis or ETosis, has been observed in various eosinophilic inflammatory diseases and is associated with disease severity.^6,8^ Through this process, the disruption of plasma and nuclear membranes results in the release of intact granules and net-like nucleus-originated DNA, known as eosinophil extracellular traps (EETs), into the surrounding tissue.^8,9^ Therefore, detecting extracellular granules is clinically important as collateral evidence of eosinophilic inflammation. However, delineating the granules with HE staining is often challenging.

Direct fast scarlet (DFS) and Congo red (CR), which are commonly used to stain amyloid fibres in diagnosing cardiac amyloidosis,^10^ are familiar to cardiologists. Previous studies have also reported enhanced visualisation of eosinophils with DFS and CR staining.^11–14^ However, whether these stains specifically mark eosinophils, and their cell-free granules has not been conclusively proven. Therefore, this study aimed to confirm whether DFS and CR can stain eosinophil granules in eosinophils and interstitial spaces and demonstrate their advantages over HE.

## METHODS

### Patients and samples

This study used endomyocardial biopsy samples obtained at the Niigata University Medical and Dental Hospital between September 2019 and July 2023. Written informed consent was obtained from each patient or their family member before the biopsy. We examined samples from six patients diagnosed with eosinophil-related heart disease and six patients diagnosed with lymphocytic myocarditis. The diagnoses of the six patients with eosinophil-related heart diseases included eosinophilic myocarditis (three patients), Loeffler endocarditis (one patient), giant cell myocarditis (one patient), and eosinophilic granulomatous polyangiitis (one patient).

### Staining procedures and image capturing

Endomyocardial biopsy samples were fixed in 10% formalin, embedded in paraffin, sliced for initial histological diagnosis, and preserved at room temperature (15–30 °C). The preserved paraffin blocks were sliced into 3-mm-thick sections, de-paraffinised, rehydrated, and stained.

#### HE staining

The samples were stained with Carrazzi’s haematoxylin solution (product number 115938; Merck Millipore CO. LTD, Hessen, Germany) for 10–30 min, rinsed in distilled water, dipped in acid alcohol (1% HCl in 70% ethanol) for a few seconds, rinsed in tap water, stained with eosin solution (product number 32042; Muto Pure Chemicals CO. LTD, Tokyo, Japan) for 2 min, sequentially transferred to 100% ethanol, cleared in xylene, and mounted.

#### DFS staining

Samples were stained with DFS solution (product number 21422; Muto Pure Chemicals CO. LTD, Tokyo, Japan) at 50 °C for 30 min, rinsed in distilled water, dipped in 80% ethanol containing 0.001% NaOH and 2% NaCl, rinsed in distilled water, stained with Mayer’s haematoxylin (made from Product number 115938; Merck Millipore CO. LTD, Hessen, Germany) for 5 min, rinsed in tap water, sequentially transferred to 100% ethanol, cleared in xylene, and mounted.

#### CR staining

The samples were stained with Mayer’s haematoxylin for 5 min, rinsed in tap water, stained with CR solution (product number 23021; Muto Pure Chemicals CO. LTD, Tokyo, Japan) for 30 min, rinsed in distilled water, dipped in 80% ethanol containing 0.001% NaOH and 2% NaCl, rinsed in distilled water, sequentially transferred to 100% ethanol, cleared in xylene, and mounted.

#### Image capturing by light microscopy

Images were captured as Tag Image File Format (TIFF) files using a microscope (Olympus BX60, Tokyo, Japan) with 60× objective lenses, a camera (Olympus DP72, Tokyo, Japan), and imaging software (Olympus DP2-BSW, Tokyo, Japan).

#### Immunofluorescence and subsequent HE, DFS, and CR staining

Eosinophils were identified by immunofluorescence in the presence of galectin-10 positivity and multiple MBP-positive granules in the cytoplasm.^7,15^ De-paraffinised sections were subjected to antigen retrieval using 0.1% proteinase K at room temperature (15–30 °C) for 6 min. The samples were incubated with the following primary antibodies: 10 μg/mL rabbit anti-human MBP (supplied by Dr. Hirohito Kita, Mayo Clinic, Scottsdale, AZ, US) for 30 min at 37 °C, and mouse anti-galectin-10 antibody in a 1:50 dilution (B-F42) (ab27417; Abcam, Cambridge, UK) for 90 min at room temperature (15–30 °C). Subsequently, the samples were incubated with Alexa Fluor 488-conjugated goat anti-mouse immunoglobulin G (IgG) (1:200 dilution; A11001; Life Technologies, CA, US), Alexa Fluor 594-conjugated goat anti-rabbit IgG (1:200 dilution; A11072; Life Technologies), and Hoechst 33342 (1:5,000 dilution; H3570; Invitrogen, MA, US) for 30 min at room temperature (15–30 °C). Samples were mounted using a Prolong Diamond (Life Technologies), and images were obtained using an LSM 980 confocal microscope (Carl Zeiss, Baden-Württemberg, Germany). After capturing immunofluorescence images, the coverslips were removed, and the samples were stained with HE, DFS, or CR. The same parts captured by immunofluorescence were recaptured using the same light microscope to elucidate whether the MBP-positive granules were stained red by HE, DFS, and CR without excess or deficiency.

### Identification of eosinophil granules by extracting red colour using Image-J

The TIFF files of HE-, DFS-, and CR-stained serial sections were captured using ImageJ software (https://imagej.net/ij/).^16^ Images were split into red, green, and blue channels, and the red channels were processed by subtracting the blue channels. The red intensity of eosinophils or their granules was compared with that of cardiomyocytes and interstitial tissues. The red intensity images were adjusted using a threshold window to selectively extract eosinophil granules while minimising background interference from cardiomyocytes and interstitial tissue.

### Counting the number of eosinophils in endomyocardial biopsy samples

Eight cardiologists evaluated the visibility of eosinophils in TIFF files of HE-stained and DFS-stained serial sections. A high-power field (using a 60× objective lens) containing several to dozens of eosinophils per field, with few background cell-free granules or degenerated eosinophils, was selected from each DFS-stained section of all 12 cases. Images of serial sections stained with DFS and HE were saved as TIFF files and colour-printed using a photocopier (Ricoh MP C6503, Japan). At this stage, the comparison of red granules in DFS with MBP-positive granules in immunofluorescence was completed, and eosinophils were defined on DFS images as spherical cells containing red granules with either a simple or bi-lobed nucleus and verified by a pathologist (H.U.).

Eight cardiologists at the Niigata University Graduate School of Medical and Dental Sciences were instructed to encircle as many eosinophils as possible with a pen within 20 s for each of the 24 prints. The DFS and HE images were arranged in an alternating, randomised order to ensure that images from the same case were not placed next to each other. Additionally, to prevent bias based on which DFS- or HE-stained sections appeared first, half of the examiners counted in one order, while the other half counted in the reverse order. A pathologist (H.U.) verified the accuracy of the circles.

### Ethics

Preserved samples were used in this study; therefore, informed consent was obtained through an opt-out option provided on the university website. The study protocol was approved by the ethics committees of Niigata University (2023-0194) and Akita University (2023-174) and was conducted in accordance with the Declaration of Helsinki.

### Patient and public involvement

Patients were not directly involved in the design or conduct of the study. However, their contributions through the provision of preserved biopsy samples were essential for addressing the study objectives. Information about the use of such samples was made publicly available on our university’s website, ensuring transparency and the opportunity for patients to opt out.

## RESULTS

### Eosinophils in HE-, DFS-, and CR-stained sections

To evaluate the appearance of eosinophils in sections stained with DFS or CR, we compared consecutive sections stained with HE, DFS, and CR of patients with various heart diseases (Figure 1). First, eosinophils were identified in HE-stained sections as small spherical cells with 1–3 lobed nuclei and small pinkish granules. Nucleus-free granules in the interstitial area were considered cell-free extracellular eosinophil granules. In consecutive sections, the distribution of reddish-orange-stained granules in DFS and CR was similar to that of the granules within eosinophils and the extracellular eosinophil granules observed in HE.

**Figure 1.**
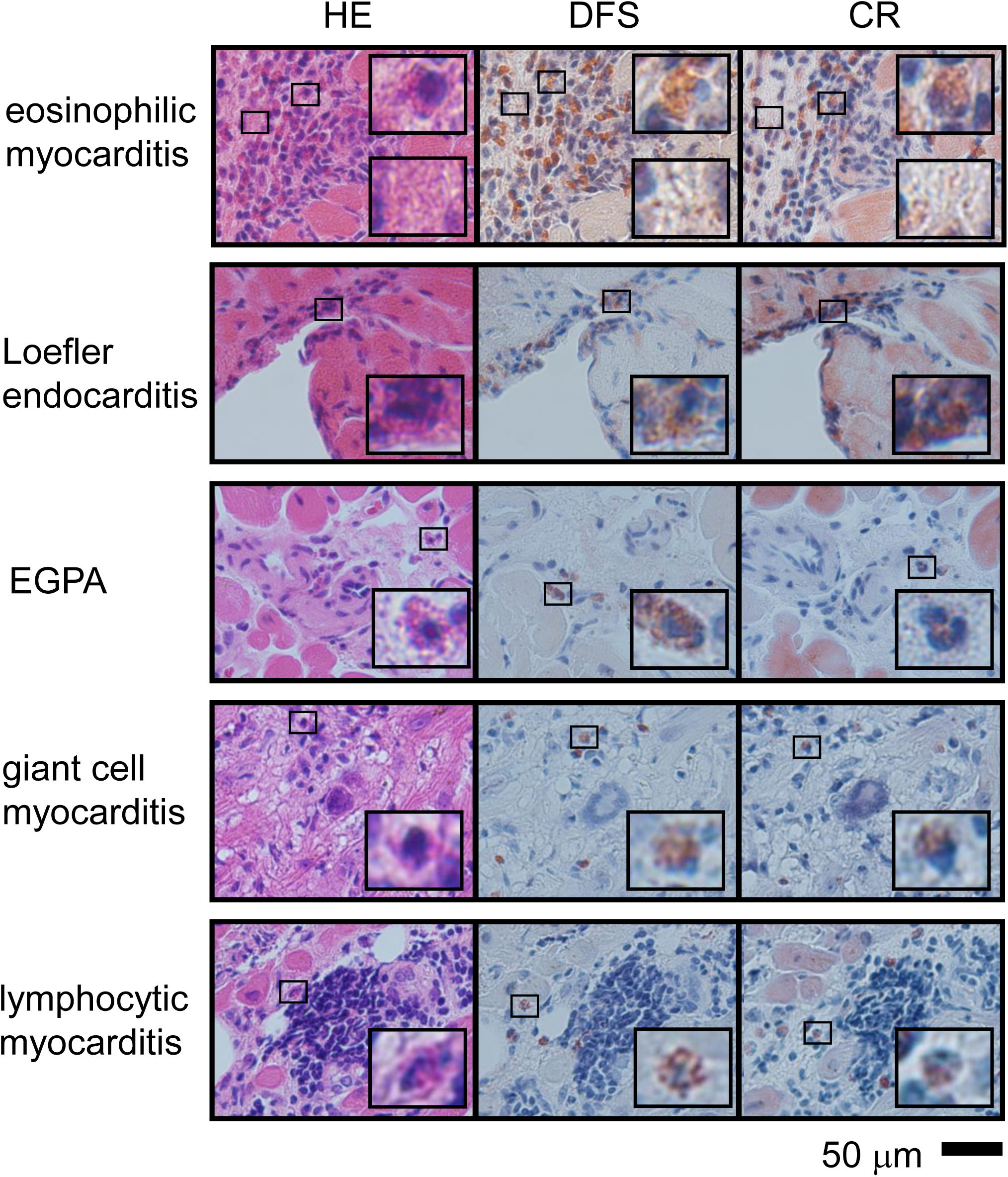
Haematoxylin-eosin-, direct fast scarlet-, and Congo red-stained endomyocardial biopsy samples from patients with various heart diseases. Insets show magnified views of eosinophils or degranulation. EGPA, eosinophilic granulomatosis with polyangiitis

Regarding visibility, while HE-stained cardiomyocytes and interstitial regions were pinkish, reducing the clarity of eosinophil granules, DFS and CR staining selectively highlighted the reddish granules with minimal staining of the interstitial regions. Particularly in DFS, cardiomyocytes remained almost unstained, making granules more readily visible than in CR.

### Comparison of immunofluorescence and histological staining

To specifically demonstrate eosinophil granules, we first identified eosinophil granules using immunofluorescence staining and subsequently counterstained the same sections with HE, DFS, or CR. Intact eosinophils, which specifically contain galectin-10 in their cytoplasm and MBP in their granules,^15,17^ were identified by immunofluorescence (Figure 2A–C, white squares). Counterstaining with HE, DFS, and CR revealed that MBP-positive granules were stained reddish, without missing any MBP-positive granules in galectin-10-positive eosinophils (Figure 2D–F, black squares).

**Figure 2.**
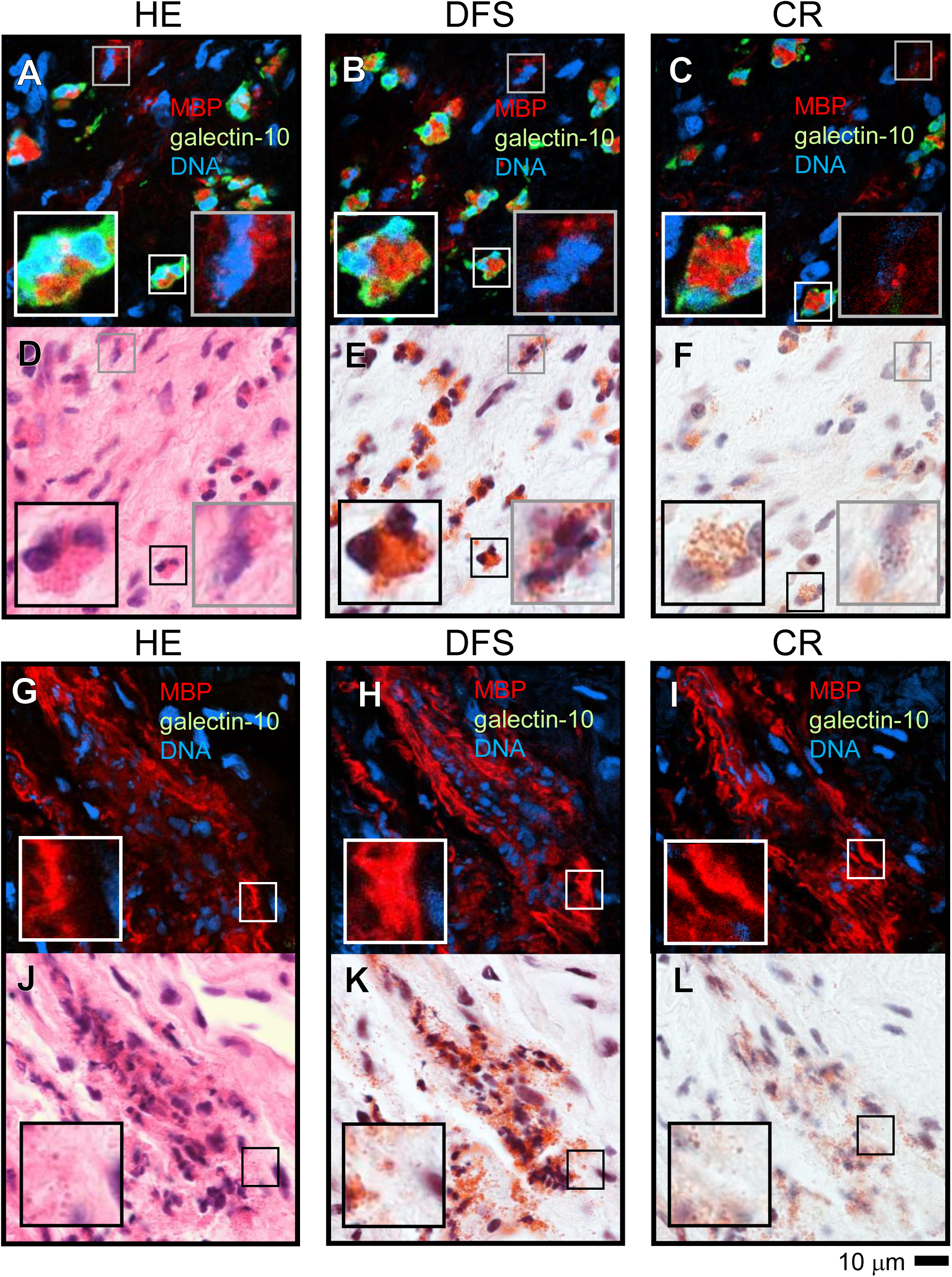
Localisation of major basic protein (MBP) by immunofluorescence and its colocalisation with red granules subsequently stained with haematoxylin-eosin (HE), direct fast scarlet (DFS), or Congo red (CR). In this representative case with eosinophilic myocarditis, eosinophils were identified using anti-MBP (red) and anti-galectin-10 (green) antibodies, and DNA is shown in blue (A–C, G–I, white squares). The same sections were subsequently stained by HE (D and J), DFS (E and K), or CR (F and L). Notably, MBP-positive granules within galectin-10-positive intact eosinophils coincide with red granules in HE, DFS, and CR (A–E). MBP-positive granules without surrounding galectin-10, suggesting granules in dying eosinophils or degranulated granules, also match with red granules stained by HE, DFS, and CR (A–E, grey squares). However, in regions without galectin-10-positive eosinophils, the MBP-positive area extended beyond red granules stained by HE, DFS, and CR (G-L, white and black squares), suggesting eosinophil granule disruption and deposition of their contents.

MBP-positive cells lacking surrounding galectin-10 fluorescence with lytic nuclei were also observed in eosinophilic myocarditis tissue (Figure 2A–F, grey squares). This staining indicated typical EETs; eosinophils had lost cytoplasmic galectin-10, and nucleus-originated DNA was released with the remaining intact granules.^15,17^ These eosinophil granules from cytolytic cells were stained with HE, DFS, and CR.

In highly damaged eosinophilic myocarditis lesions, galectin-10-positive cells were absent, whereas diffuse or linear fluorescence patterns were observed using an anti-MBP antibody (Figure 2G–I). Netlike DNA was observed in these lesions, indicating the presence of EETs. Counterstaining with HE, DFS, and CR detected reddish granules; however, it did not reproduce diffuse or linear patterns (Figure 2J–L). These results indicate that DFS and CR are as effective as HE in detecting intact eosinophil granules; however, these staining techniques are unable to detect tissue deposition of eosinophil granule proteins after granule disruption.

### Extraction of eosinophil granules from background tissue using Image-J

To evaluate whether DFS and CR were more effective than HE in objectively recognising eosinophil granules, we assessed the feasibility of selective eosinophil granule extraction through red colour segmentation using ImageJ software. Representative images of red colour extraction from the HE, DFS, and CR images are shown in Figure 3.

**Figure 3.**
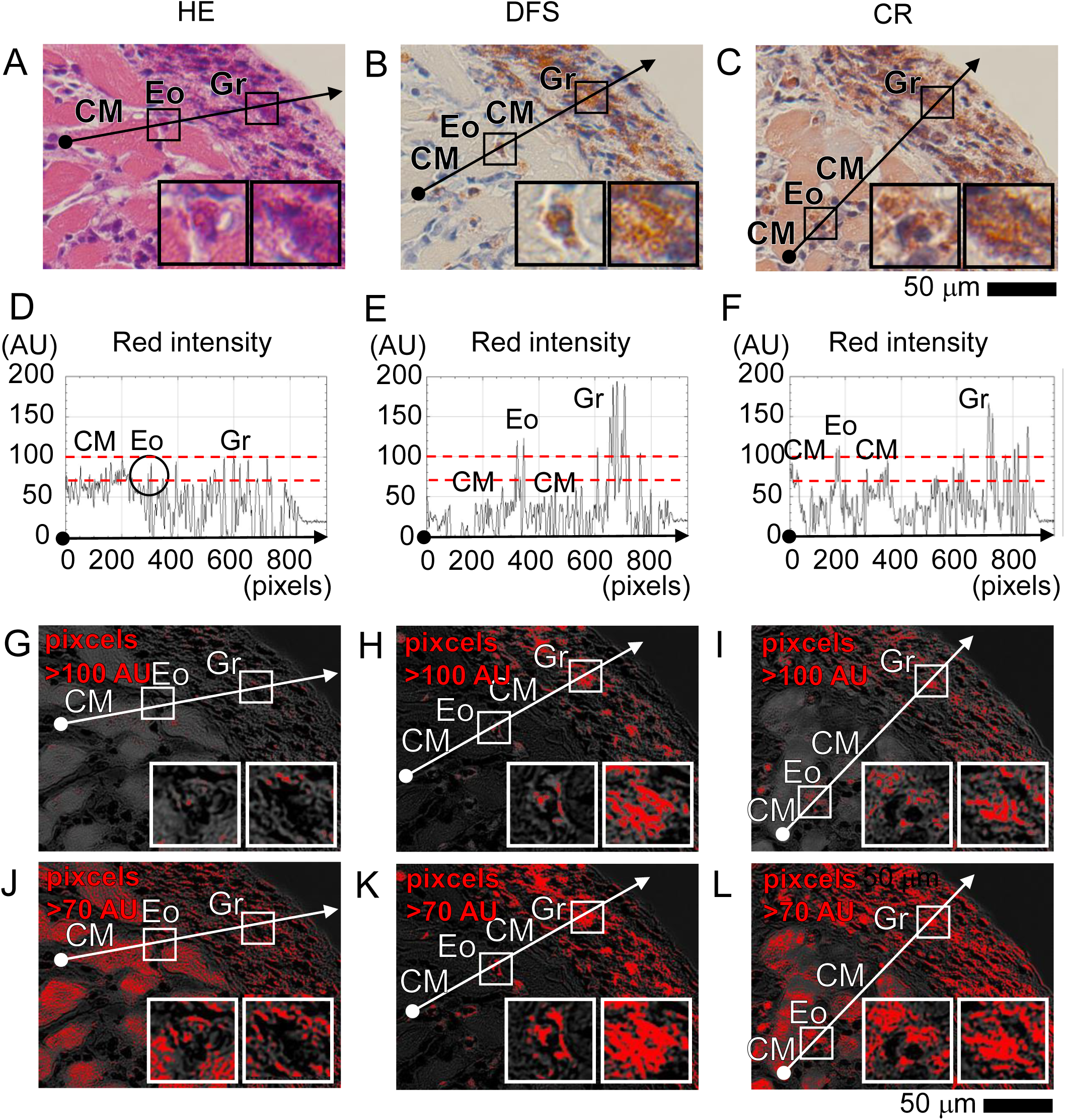
Representative images showing the advantage of staining by direct fast scarlet (DFS) over haematoxylin-eosin (HE) and Congo red (CR) in identifying eosinophils and their granules. Panels A, B, and C are serial sections stained by HE, DFS, and CR. The intensity of red colour on the arrows in A, B, and C are shown in graphs D, E, and F. The red intensities of cardiomyocyte (CM), eosinophil (Eo), and degranulated eosinophil granules (Gr) were similar in HE staining (D). In DFS staining, the red intensity of CM was lower than that of Eo and Gr (E). In CR staining, the intensity of CM was higher than that in DFS and closer to that of Eo. The red intensity of greater than 100 arbitrary unit (AU) on A, B, and C are shown as red on G, H, and I, highlighting that selective visualisation of Eo and Gr was possible with DFS and CR staining. However, in images showing red intensity greater than 70 AU (J, K, and L), only DFS could distinguish Eo from CM. Insets in each image provide magnified views of an eosinophil and cell-free granules.

In HE-stained sections, the red intensity of eosinophil granules was comparable to that of cardiomyocytes (Figure 3A, D), making it challenging to selectively extract eosinophil granules (Figure 3G, J). In contrast, DFS staining exhibited high red intensity in eosinophil granules and low red intensity in cardiomyocytes (Figure 3B, E), enabling the efficient detection of eosinophil granules both within eosinophils and in the interstitial space (Figure 3H, K). CR also allowed eosinophil granule extraction; however, the reddish hue of cardiomyocytes (Figure 3C, F) led to their overextraction when the red intensity threshold was lowered (Figure 3I, L). In summary, DFS was superior to CR in identifying eosinophil granules through red colour segmentation, whereas HE was ineffective.

### Eosinophil counting test

To demonstrate that DFS facilitates easier identification of eosinophils than HE by non-pathological clinicians, cardiologists without pathology training performed the eosinophil counts. Figure 4A and 4B present cases of eosinophilic myocarditis. The green circles in panels A and B indicate the 13 eosinophils pre-counted in a DFS-stained image. Across 12 DFS-stained images, 113 eosinophils were pre-counted. Panels C and E display representative images of printed serial sections used for the test, with panel C showing DFS staining and panel E showing HE staining. Panels D and F present magnified views of the regions containing eosinophil clusters from panels C and E, respectively

**Figure 4.**
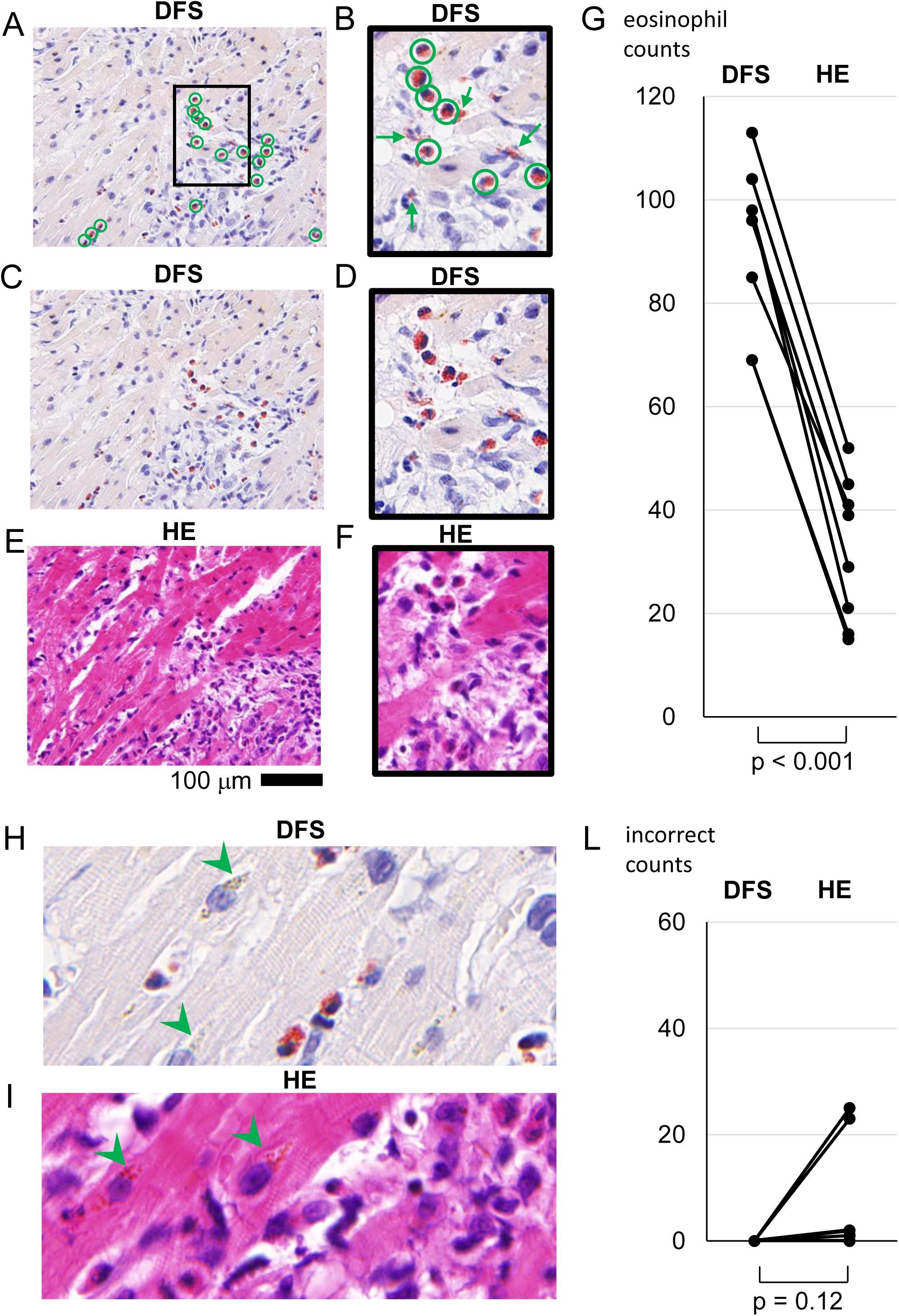
Representative images used for eosinophil counting test and the results. Green circles in panels A and B indicate cells pre-identified as eosinophils before the counting test. Green arrows in panel B point to excluded red granules due to the absence of a nucleus or a non-round shape. Panels C and E show the images used for the eosinophil counting test, while panels D and F present magnified portions of panels C and E, respectively. Panel G displays a comparison of eosinophil counts by each cardiologist in 12 direct fast scarlet (DFS)-stained and 12 haematoxylin-eosin (HE)-stained images. All eight cardiologists identified more eosinophils in DFS-stained images (87 ± 22) than in HE-stained serial section images (32 ± 14) out of 113 pre-identified eosinophils. Panels H and I demonstrate representative instances of incorrect counts. Some perinuclear granules in cardiomyocytes (CMs) were stained reddish even with DFS staining (H, arrowheads). In some HE-stained CMs, perinuclear granules (I, arrowheads) were stained as red as eosinophilic granules, leading to incorrect counts. L shows the number of incorrect counts made by each cardiologist.

When counting the pre-counted 113 eosinophils across the 12 images, each evaluated within 20 s, eight cardiologists counted an average of 87 ± 22 eosinophils on DFS- and 32 ± 14 on HE-stained serial sections, with significantly higher counts on DFS (Figure 4G). In contrast, eosinophil misidentification occurred with HE staining, in which the cardiomyocytes were occasionally mistaken for eosinophils. Some perinuclear granules in the cardiomyocytes displayed a red hue, even in DFS-stained sections (Figure 4H arrowheads). The HE-stained sections exhibited a stronger red colouration of the perinuclear granules (Figure 4I arrowheads), leading to misidentification, particularly by inexperienced individuals. The number of miscounts was reduced among some cardiologists (Figure 4L); however, this reduction was not statistically significant.

## DISCUSSION

Meaningful interpretation of findings in endomyocardial biopsy is achieved only when clinicians or cardiologists integrate them with information on the patient’s history and the results of multiple diagnostic examinations.^3–5^ The advantage of using DFS and CR staining for eosinophil detection shown in this study lies in their high visibility, which enables easy recognition, including its released granules, at a glance, even by a non-pathologist. Notably, DFS shows the potential for machine detection through colour extraction. The high visibility of these stains has been previously noted using serial sections in eosinophilic myocarditis^3^ and eosinophilic inflammation of other organs; ^11,12^ however, no study has comprehensively demonstrated these findings across various heart diseases. Furthermore, eosinophil MBP-positive granules are stained red with HE in the same section; ^18^ however, our study is the first to show that MBP-positive granules are stained by DFS and CR, providing direct evidence that eosinophil granules, even if present extracellularly, are recognisable using DFS and CR staining.

Alternative methods to enhance eosinophil detectability over that of HE include immunofluorescence or immunohistochemistry. However, these approaches are time- and cost-consuming, and require specific antibodies, equipment, and technical expertise. In contrast, DFS and CR are simple and cost-effective methods that can be implemented globally and are already widely used to diagnose cardiac amyloidosis.^9,18^ Among other staining techniques, chromotrope 2R resembles DFS in molecular structure and showed a similar staining pattern to ours with DFS.^14^ In that report, CR was also examined, resulting in reddish staining of surrounding tissues, and DFS offered superior visibility, consistent with the findings of our study.

One of the clinical issues concerning eosinophil counting is the lack of clear criteria for diagnosing eosinophilic myocarditis, such as the number of eosinophils per microscopic field or the percentage of eosinophils in infiltrating cells, which sometimes leads to uncertainty in discrimination from lymphocytic myocarditis,^3,20^ which usually does not require immunosuppressive therapy.^3^ In the current study, DFS facilitated easier eosinophil counting in myocardial tissue by improving the signal-to-noise ratio and demonstrated potential for automated counting through red colour extraction. As a reliable detection method, DFS may lead to the establishment of numerical diagnostic criteria and the determination of therapeutic indications for eosinophilic myocarditis and other eosinophil-related heart diseases.

Eosinophil granules contain highly cytotoxic cationic proteins.^6,21^ Therefore, once eosinophil granules are detected, assessing both their release and the subsequent release of specific granule proteins into the interstitial space is important. Galectin-10 is expressed in the cytoplasm of intact eosinophils, disappears during ETosis, and exerts toxicity towards microorganisms or self-tissues.^15^ In the present study, we observed that MBP-positive granules were detectable with DFS and CR in all phases: in intact eosinophils, when surrounding galectin-10-negative dead eosinophils, and after being scattered in the interstitial space. However, in the state where MBP is diffusely deposited, DFS and CR fail to stain these components red, suggesting that they are unsuitable for detecting granule content after granule disruption. This finding was reported using HE in 2011,^22^ and recent reviews have reported similar observations in other organs,^6^ although no subsequent studies have addressed this in the cardiac tissue. The disappearance of red staining in DFS and CR is an important characteristic when assessing the stage of eosinophilic myocardial injury and considering the treatment strategy because immunosuppressive therapy may not be necessary once eosinophils and their granules disappear, even if the contents of eosinophil granules remain deposited.

Recent studies have suggested that eosinophils have cardioprotective roles in myocardial infarction, heart failure, and hypertrophy.^23–25^ The ability to detect eosinophils in cardiac diseases through myocardial biopsy or autopsy, even when the number of eosinophils is low, could enhance our understanding of disease mechanisms and guide treatment. Therefore, effective detection of eosinophils using DFS is considered a valuable tool.

### Study Limitations

This study has some limitations. First, it included a limited number of biopsy cases from a single hospital, which might affect the generalisability of the findings. Second, we did not evaluate the utility of DFS and CR in frozen sections of endomyocardial biopsies obtained during a clinical emergency. Third, the biopsy samples analysed were limited to patients with eosinophil-infiltrating diseases or lymphocytic myocarditis. Validation in other major cardiac conditions, such as acute myocardial infarction, is required because neutrophils may also be faintly stained red by DFS and CR.^12,26^ Finally, this study focused on the presence of eosinophilic granules without addressing their functional roles. The detection of cell-free extracellular eosinophil granules may be associated with the role of eosinophils, whether they are protective or contribute to tissue damage; however, further investigation is required to substantiate this.

## CONCLUSIONS

This study demonstrated that eosinophils and eosinophil granules in the myocardial tissue were more readily detected with DFS and CR than with HE, with DFS showing particular superiority. Importantly, DFS and CR did not stain the deposited granule proteins in the tissue. DFS may pave the way for a deeper understanding of the role of eosinophils in heart disease.

## Data Availability

All data produced in the present study are available upon reasonable request to the authors

## Acknowledgements

The authors thank Noriko Tan for her exceptional technical assistance, particularly in acquiring high-quality immunofluorescence images.

## Contributors

TKu: Clinical data collection, histological analysis, ImageJ software analysis, and manuscript writing

TA: Staining, immunofluorescent, histological analysis

TKa: Study conception, image-J analysis, manuscript writing and editing SU: Funding, Study conception, manuscript editing, supervision

HU: Pathological confirmation

SS, SH, HI, KT, YS, HT, RS, HK, SF, TT: Endomyocardial biopsy, eosinophil count test, critical discussion

TI: Funding, manuscript editing, supervision

## Patient consent for publication

Not required.

## Ethics approval

The study protocol was approved by the ethics committees of Niigata University (2023-0194) and Akita University (2023-174) and was conducted in accordance with the guidelines of the Declaration of Helsinki.

